# Evaluating the effectiveness of the HPV vaccination programme in England, using regression discontinuity design

**DOI:** 10.1101/2024.04.19.24306076

**Authors:** Isobel L. Ward, Charlotte R. Bermingham, Kate Soldan, Vahé Nafilyan

## Abstract

In England, the National human papillomavirus (HPV) immunisation programme was introduced in 2008 to prevent cervical cancer. Girls aged 12 to 13 were offered routine vaccination, and those aged 14 to 18 years in 2008 were offered ‘catch-up’ vaccination. Using Census 2011, Hospital Episode Statistics and mortality data for the population of England, we exploit the cut-off in eligibility and apply a regression discontinuity design (RDD) to assess the impact of HPV vaccination on cervical disease. Vaccination reduced the incidence of cervical dysplasia and cancer diagnoses by 31% and 75% respectively at ages 23 to 30 years in girls offered catch-up vaccination at ages 17 to 18 years compared to those who were just above the eligibility age for the catch-up vaccination, with a clear discontinuity. Reductions continued amongst girls offered routine vaccination. These estimates obtained with a quasi-experimental approach are similar to vaccine effectiveness estimates based on more traditional approaches. This novel approach provides further evidence of the HPV vaccination programme reducing adverse cervical outcomes in young women and could be used for future studies to evaluate major changes in HPV vaccination policy and for studies of longer-term outcomes including other cancers and deaths.

## Introduction

Human papillomavirus (HPV) is a very common virus which infects squamous epithelial cells in both men and women. The majority of HPV infections are transient and result in no clinical problems, however persistent infection by a ‘high-risk’ HPV type is the cause of most cervical pre-cancerous and cancerous lesions. In high-income countries the incidence and mortality of cervical cancer has been decreasing over the past 30 years following the introduction of formal screening programmes. In the UK, an estimated 1 in 130 females will be diagnosed with cervical cancer in their lifetime [1].

The National HPV immunisation Programme was introduced in 2008 [2] to reduce the incidence of cervical cancer in women. From 1^st^ September 2008, girls in school year 8 (ages 12 to 13), born September 1995 to August 1996, were the first birth cohort offered routine vaccination [3]. In addition to this routine programme, a ‘catch-up’ programme was also launched in 2008 to offer girls aged 13 to 18 years of age (born September 1990 to August 1995) HPV vaccination. In 2008/09 catch-up vaccination was offered to females aged 17 to 18 years and in 2009/10 females aged 14 to 18 in four catch up birth cohorts [2] (**Supplementary** Figure 1). The bivalent HPV vaccine (Cercarix ©) provides direct protection against the two most common high-risk types of HPV (16 and 18), responsible for over 70% of all cervical cancers in England [4], and was the vaccine used in the national programme for the first four years. The programme switched in September 2012 to using a quadrivalent vaccine (Gardasil ©) [5] which provides additional direct protection against the two low-risk types (6 and 11) that cause over 90% of anogenital warts [6].

In clinical trials both these HPV vaccines were over 90% effective at preventing pre-cancers of the cervix caused by HPV types 16 and 18 in young women [7], [8]. Recently published work assessing the impact of the HPV vaccination programme on cervical cancer in England and Scotland has shown significant reductions in the number of pre-cancers (grade 3 cervical intraepithelial neoplasia (CIN3)) and cancer diagnoses [9], [10]. The majority of cases of cervical dysplasia are caused by persistent infection of an HPV virus. Falcaro et al. (2021) found that girls who were offered HPV vaccination at 12 to 13 years had a reduction of 87% in incidence of cervical cancer and those offered vaccination at 16 to 18 years had a 34% reduction, in data to mid-2019, up to age 28 years [9]. Palmer et al (2024) analysed Scottish cervical screening data linked to cancer registry, immunization and deprivation data, to mid-2020, and found no cancer cases in women immunized though the routine programme (aged 12 to 13 years) and an adjusted vaccine effectiveness for cancers of 74% in fully-vaccinated women in the catch-up programme [10].

Assessment of the impact of vaccination in real world conditions is essential information for policy makers and clinicians; especially for longer term outcomes not included in clinical trials, such as cervical cancer, and for assessing the impact outside of the controlled conditions of a clinical trial and when applied to the general population. However, estimating vaccine effectiveness based on real-world data is challenging, as unvaccinated individuals may differ from vaccinated individuals in ways that are not easy to measure. For example, vaccinated individuals may be more likely to undergo screening due to health seeking behaviours and therefore may be more likely to receive a cervical dysplasia and/or early cancer diagnosis and less likely to receive a late cancer diagnosis. Conversely, healthier individuals may be more likely to be vaccinated, and may have lower risk of HPV and related cancer due to behavioural factors. However, determining variables to adjust for these differences is challenging due to the many factors involved and lack of a clear measurement of ‘health’, which can result in unmeasured confounding. In addition, temporal trends may have changed the outcome without the intervention.

Here, we used data from the Public Health Data Asset, a population-level dataset which combines the 2011 Census linked to Hospital Episode Statistics and Office for National Statistics (ONS) death registrations data [11] to create a linked dataset for the population of England to analyse cervical outcomes for women in England who were eligible for the first two years of catch-up HPV vaccination, and for three older birth cohorts who were not. We use a regression discontinuity design to estimate the effectiveness of the HPV vaccine in preventing cervical disease.

Observational studies produce estimates of vaccination impact that are not biased by measured confounding factors, by adjusting for these statistically using cohort study or case-control study designs but confounding by unmeasured factors may remain [12]. Population-level impact studies, such as ours using a RDD approach offer a complimentary technique to these studies, producing estimates of vaccination impact that are not biased by unmeasured factors that may affect uptake and cause confounding.

## Methods

### Overview of the approach

We used a regression discontinuity design to estimate the effect of the HPV vaccine on a range of outcomes. Regression discontinuity design is a quasi-experimental technique used in situations where individuals are assigned to treatment based on a cut-off (here academic year of birth), where the change (or not) in the outcome at the eligibility cut-off is determined to provide an estimate for the causal effect of eligibility for treatment [13], [14]. The change in treatment coverage at the cut-off is used to estimate the causal effect of treatment.

Regression discontinuity designs can be applied to estimate vaccine effectiveness by exploiting sudden differences in vaccination coverage, such as due to changes in eligibility for vaccination [15].

In the main analysis, we compared outcomes in the cohorts that were part of the catch-up vaccination programme to cohorts that did not benefit from the programme. In our secondary analysis, we compared outcomes in the cohorts that were part of routine vaccination and cohorts that were part of the routine vaccination programme.

### Study data

We used data from the Office for National Statistics (ONS) Public Health Data Asset (PHDA), a linked dataset combining the 2011 Census, Office for National Statistics death registrations and electronic health records [11]. Diagnosis outcomes were sourced from Hospital Episode Statistics (HES). The HES dataset includes finished hospital episodes that ended from 1 April 2009 to 30 August 2022. The mortality dataset includes all deaths that occurred up to 20 December 2023 that were registered between 7 January 2011 and 20 December 2023.

HPV vaccination coverage estimates were obtained from UKHSA based on administered vaccinations data and serology prevalence estimates [2], [16]. Ethical approval was obtained from the National Statistician’s Data Ethics Advisory Committee (NSDEC).

### Study population

For the main analysis, we compare women who we not eligible for vaccination to those who were eligible for catch-up vaccination. The eligibility cut-off is at the birth month where catch-up vaccination was first introduced. We included women who were in the three birth cohorts just above the age eligible for HPV vaccination (birth dates 1 September 1987 to 31 August 1990) or were in the oldest two birth cohorts eligible for catch-up vaccination (birth dates 1 September 1990 to 31 August 1992), to assess the impact of the catch-up vaccination programme (**Supplementary Figure 2a**). For the secondary analysis, we compare women who were eligible for catch-up vaccination to those eligible for routine vaccination. The eligibility cut-off is at the birth month at which catch-up vaccination switched to routine vaccination. We included women who were in the youngest three birth cohorts eligible for catch-up vaccination (birth dates 1 September 1992 to 31 August 1995) or were in the first two birth cohorts eligible for routine vaccination (birth dates 1 September 1995 to 31 August 1997) (**Supplementary Figure 2b**).

For both analyses, all women were enumerated in the 2011 Census, usual resident in England on 2011 Census day (27 March 2011) and were alive at the start of follow up (age 23 years for the main analysis and 19 years for the secondary analysis).

### Outcome

Two cervical diagnosis outcomes were analysed: 1) diagnosis of cervical dysplasia 2) diagnosis of cervical cancer. Diagnoses of cervical cancer or cervical dysplasia (ICD N870-N879) were identified as a hospital episode with primary or secondary cause of admission coded as these causes in the HES Admitted Patient Care (APC) dataset (**Supplementary Table 3**). We also carried out a supplementary analysis where the outcome was death involving cervical cancer, identified as a death with underlying or contributory cause of death with ICD10 code corresponding to cervical cancer (C530-C539) in ONS death registrations.

Diagnosis outcomes were identified over a 7 year range from age 23 to 30 years in the main analysis and over a six year range from age 19 to 25 years in the secondary analysis. The age ranges allowed 7 years of follow-up for all individuals in the first analysis and 6 years of follow-up for the second analysis. The follow up age ranges were determined by the limits of the HES APC data (minimum episode data 1 April 2009 and maximum episode data 31 August 2022) and mortality data (deaths up to 31 August 2022) available at time of analysis for the included birth cohorts (Supplementary Figure 2). The age range for the main analysis covered two cervical screening invitations for most women in England who are invited for their first cervical screening 6 months before their 25^th^ birthday and for a follow up screening 3 years after their first. The age range for the secondary analysis covered the first invitation for cervical screening only.

The start dates of all recorded hospital episodes with the relevant diagnosis code present were recorded, and the outcome was a binary variable for each individual representing that at least one episode occurred within the follow up period (23-30 for the older catch-up versus non-vaccinated, or 19-25 for the routine versus younger catchup (**Supplementary** Figure 2).

### Intervention

HPV vaccination rates by academic year of birth for vaccinated cohorts were obtained from the UK Health Security Agency (UKHSA) based on administered vaccinations data and serology prevalence estimates [2], [16]. We used the estimates for having two doses in our analysis. Since 2014 the Joint Committee on Vaccination and Immunisation (JCVI) have been recommending a two dose HPV vaccination schedule, and since 2023 a one dose schedule.

We derived estimates of coverage by month of birth by assuming constant vaccination coverage across months within an academic year of birth. No published vaccination coverage data was available for birth cohorts who were not eligible for vaccination, however it is unlikely to have be of any substantial importance at population level due to the cost of private vaccination [17]. The coverage was therefore estimated to be a nominal 0.01% for all cohorts not eligible for vaccination. Estimates of coverage per-dose for each birth cohort are provided in Supplementary Tables 2 and 3.

### Eligibility

Eligibility for catch-up vaccination in the main analysis and routine vaccination in the secondary analysis were determined by the month of birth. For the main analysis, women born before 1 September 1990 were ineligible for any HPV vaccination (the non-vaccine eligible study cohort) and those born after this date were eligible for catch-up vaccination (the older catch-up study cohort, aged 17 to 18 at vaccination) [2]. For the secondary analysis, girls were eligible for the routine vaccination (the routine study cohort) if they were born after 1 September 1995 and those born before this date were eligible for catch-up vaccination at ages 14 to 17 (the younger catch-up study cohort). Date of birth was grouped into birth month to assess our outcomes.

### Statistical Analyses

We calculated diagnosis and mortality rates for women aged 23 to 30 between September 2010 and August 2022 (no vaccination versus older catch-up) over the 7-year-follow-up by month of birth. We combined these rates with HPV vaccination rates by month of birth.

We used these data to estimate the effect of the HPV immunisation programme and derive an estimate of the impact of HPV vaccination using a RDD approach. First, to estimate the overall effect of the effect of the HPV immunisation programme on mortality and diagnosis rates, we used a sharp RDD design, with the exposure being eligibility to vaccination and the model being adjusted for month of birth linearly, allowing for different slope in the eligible and non-eligible cohorts. Second, we used a fuzzy RDD, which takes into account HPV vaccination coverage by birth month, to obtain an estimate of the impact of receiving an HPV vaccination on mortality rates and diagnosis rates. By scaling the effect of eligibility by the difference in vaccination coverage, the fuzzy RDD gives us an estimate of the Local Average Treatment Effect (LATE), that is the effect of vaccination on these outcomes among girls who wouldn’t have been vaccinated in the absence of the programme.

Third, to calculate the impact of vaccination, we fitted the same model but used the log odds of cervical outcomes as the dependent variable instead of the rates of cervical outcomes. This allowed us to estimate the odds ratio of being eligible for vaccination and to derive the vaccine effectiveness as one minus the odds ratio.

The parameters of interest were estimated non parametrically using local polynomial estimators [18], as implemented by the R package rdrobust. In the main specifications, we used a uniform kernel, and the effect of the running variable was assumed to be linear on both side of the cut-off, but the slopes were allowed to be different. We used a bandwidth of 36 months before the cut-off and 24 months after the cut-off. As the RDD is fitted to the rates of outcomes over the follow-up period, the model fitting was weighted by the number of girls in each month.

We also reproduced the results for the secondary analysis of women vaccinated as part of both the younger catch-up and routine study cohorts using a follow up of 6-years from ages 19 to 25 years between September 2011 and August 2022.

The validity of the RDD rests on the assumption that in the absence of the intervention there would have been no discontinuity in the outcome. Whilst this assumption cannot be tested directly, we checked for continuity across the eligibility cut-off in the distribution by quintiles of IMD and ethnicity. We also estimated the discontinuity for death not caused by cancer as this should not be affected by vaccination. To ensure that the discontinuity in rates of cervical outcomes was caused by eligibility for the vaccine, we estimated discontinuity in outcomes at different cut-offs at the eligibility point (months 34 to 38).

All data engineering was done in Python 3.6.8 and statistical analyses were done in R version 3.5.1. The RDD was estimated using the rdrobust package.

### Patient and Public involvement

No members of the public were involved with this analysis.

## Results

In our main analysis we have a total of 1,445,512 women born between 1 September 1987and 31 August 1992 **(Table 1)**. The proportion of different ethnic groups and proportion belonging to different IMD groups is similar between the older catch-up and the non-vaccination eligible cohorts. **(Table 2)**. The estimated vaccination rate increased from 0.01% to over 55% following the introduction of the HPV vaccination programme **(Supplementary Table 2)**.

**Table 1:** Total number of individuals in the study population by birth cohort and vaccination group.

| Analysis | Birth cohort | Vaccination group | Total (n) |
| --- | --- | --- | --- |
| Main analysis | 1 Sept 1987 - 31 Aug 1988 | Non vaccine eligible | 309,536 |
|  | 1 Sept 1988 - 31 Aug 1989 | Non vaccine eligible | 291,784 |
|  | 1 Sept 1989 - 31 Aug 1990 | Non vaccine eligible | 281,293 |
|  | 1 Sept 1990 - 31 Aug 1991 | Older catch-up vaccination | 281,030 |
|  | 1 Sept 1991 - 31 Aug 1992 | Older catch-up vaccination | 281,869 |
| Secondary analysis | 1 Sept 1992 - 31 Aug 1993 | Younger catch-up vaccination | 291,192 |
|  | 1 Sept 1993 - 31 Aug 1994 | Younger catch-up vaccination | 289,786 |
|  | 1 Sept 1994 - 31 Aug 1995 | Younger catch-up vaccination | 281,799 |
|  | 1 Sept 1995 - 31 Aug 1996 | Routine vaccination | 283,604 |
|  | 1 Sept 1996 - 31 Aug 1997 | Routine vaccination | 283,105 |

**Table 2:** Characteristics of non-vaccinated and older catch-up cohorts used in the main analysis and the routine and younger catch-up cohorts used in the secondary analysis.

| Characteristic | Level | Non vaccine eligible |  | Older catch-up vaccination |  |
| --- | --- | --- | --- | --- | --- |
|  |  | Total (n) | Proportion | Total (n) | Proportion |
| Ethnicity | Asian | 86829 | 9.8 | 48417 | 8.6 |
|  | Black | 30704 | 3.5 | 20055 | 3.6 |
|  | Mixed | 25623 | 2.9 | 18077 | 3.2 |
|  | Other ethnic group | 7975 | 0.9 | 4795 | 0.9 |
|  | White | 731482 | 82.9 | 471555 | 83.8 |
| Index of Multiple Deprivation | 1 | 221581 | 25.1 | 132939 | 23.6 |
|  | 2 | 206638 | 23.4 | 125081 | 22.2 |
|  | 3 | 175092 | 19.8 | 110219 | 19.6 |
|  | 4 | 149566 | 16.9 | 101964 | 18.1 |
|  | 5 | 129736 | 14.7 | 92696 | 16.5 |
| Characteristic | Level | Younger catch-up vaccination |  | Routine vaccination |  |
|  |  | Total (n) | Proportion | Total (n) | Proportion |
| Ethnicity | Asian | 73281 | 8.5 | 49332 | 8.7 |
|  | Black | 34584 | 4.0 | 23465 | 4.1 |
|  | Mixed | 30337 | 3.5 | 22117 | 3.9 |
|  | Other ethnic group | 7084 | 0.8 | 4949 | 0.9 |
|  | White | 717491 | 83.2 | 466846 | 82.4 |
| Index of Multiple Deprivation | 1 | 180933 | 21.0 | 120335 | 21.2 |
|  | 2 | 165425 | 19.2 | 108763 | 19.2 |
|  | 3 | 163409 | 18.9 | 107228 | 18.9 |
|  | 4 | 168864 | 19.6 | 110162 | 19.4 |
|  | 5 | 184146 | 21.3 | 120221 | 21.2 |

The 7-year rates of diagnosis of cervical dysplasia and by birth month are shown in **Figure 1 (Supplementary Table 4)**. Crude 7-year rates in outcomes by study cohort are reported in **Table 3**. Overall, all cervical outcomes had lower rates of occurrence over the 7-year follow up per 100,000 persons in girls who were eligible for vaccination as part of the older catch-up programme compared to those not eligible for vaccination.

**Figure 1:**
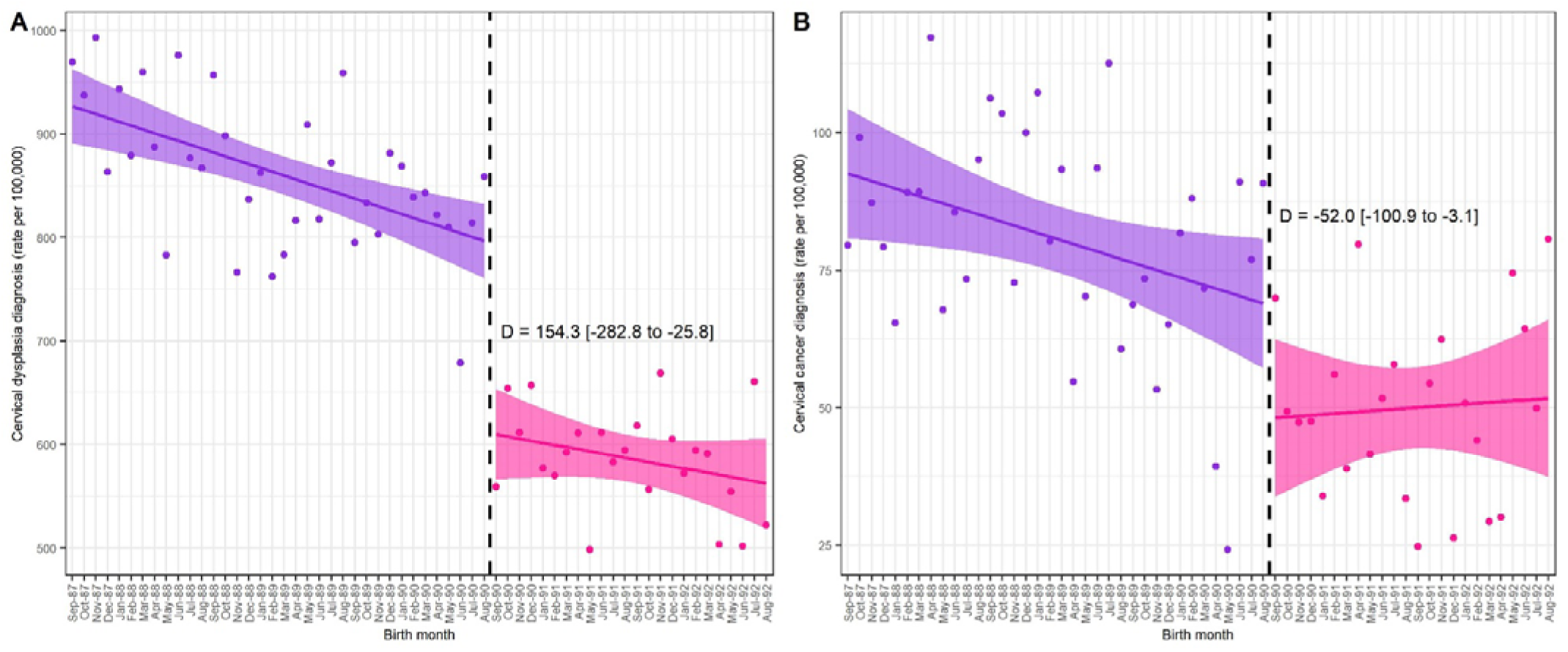
Rate of cervical dysplasia (A) and cervical cancer (B) outcomes across a 7-year follow up period in girls who were eligible in oldest 2 years of catch-up and not eligible for HPV vaccination by month of birth. Regression discontinuity design plots for those eligible for HPV vaccination as part of a catch-up programme (aged 17-18) versus those non-eligible. The rates for those not eligible for the HPV vaccination are shown in purple, and pink for girls eligible for HPV vaccination as part of the catch-up programme. Girls born from 1 September 1990 were eligible for the catch-up vaccination when they were ages 17 to 18 years. D is the difference in the rate of diagnosis at the discontinuity.

**Table 3:** Rates of cervical outcomes, and non-cancer outcomes for each analysis. Total counts and rates of events for the main (non-vaccine eligible versus older catch-up) and secondary analysis (younger catch-up versus routine). Rates for the main analysis are over a 7-year follow-up period, and a 6-year period for the secondary analysis.

|  | **Main analysis** |  | **Secondary analysis** |  |
| --- | --- | --- | --- | --- |
|  | **Non vaccine eligible** | **Older catch-up** | **Younger catch up** | **Routine** |
| Total (n) | 882613 | 562899 | 862777 | 566709 |
| Cervical dysplasia diagnosis (total) | 7622 | 3299 | 877 | 405 |
| Cervical cancer diagnosis (total) | 715 | 281 | 62 | 17 |
| Cervical cancer mortality (total) | 51 | 22 | 4 | 0 |
| Any other non-cancer mortality (total) | 1577 | 1076 | 1090 | 685 |
| Cervical dysplasia diagnosis (rate per 100,000) | 863.57 | 586.07 | 101.65 | 71.47 |
| Cervical cancer diagnosis (rate per 100,000) | 81.01 | 49.92 | 7.19 | 3 |
| Cervical cancer mortality (rate per 100,000) | 5.78 | 3.91 | 0.46 | 0 |
| Death not caused by cancer (rate per 100,000) | 178.67 | 191.15 | 126.34 | 120.87 |

There was a significant discontinuity in the rate of cervical dysplasia diagnosis around the cut-off for being eligible for the catch-up programme (**Figure 1**). Being part of the older catch-up study cohort was associated with a reduction of 154 (95%CI:26-283) cases per 100,000 people, at ages 23 to 30 years. The fuzzy RDD analysis (**Table 4**) indicates that receiving the HPV vaccination at age 17 to 18 as part of the catch-up program reduced cervical dysplasia diagnoses by 260 (95%CI:44-477) cases per 100,000 people between the ages of 23 to 30 years. This corresponds to a vaccine effectiveness of 31.7% (95%CI:5.7-49.2) for the oldest girls who were vaccinated as part of the catch-up programme against cervical dysplasia.

**Table 4:** Estimates of the effect of vaccine effectiveness on cervical dysplasia and cancer diagnosis in women ages 23 to 30 offered catch-up HPV vaccination aged 16 to 18 years.

| Value | Cervical dysplasia diagnosis | Cervical cancer diagnosis | Death not caused by cancer |
| --- | --- | --- | --- |
| Discontinuity/100,000 (95% CIs) | -154.28 (-282.76, -25.79) | -52.04 (-100.94, -3.13) | -2.13 (-49.06, 44.79) |
| LATE/100,000 (95% CIs) | -260.21 (-476.92, -43.5) | -87.77 (-170.26, -5.28) | -3.6 (-82.75, 75.55) |
| p-Value | 0.02 | 0.04 | 0.93 |
| Vaccine effectiveness (%) | 31 (6, 49) | 75 (-11, 95) | 2 (-51, 37) |

For cervical cancer diagnoses, being vaccinated as part of the catch-up was associated with a reduction of 52 (95%:CI 3-101) cases per 100,000 people (**Figure 1).** The fuzzy RDD analysis (**Table 4**) indicates that receiving the HPV vaccination at age 17 to 18 as part of the catch-up program reduced cervical cancer diagnoses by 88 (95%CI:5-170) cases per 100,000 people between the ages of 19 to 25 years. This corresponds to a vaccine effectiveness of 75.4% (95%CI:11.4-94.6) for the oldest girls who were vaccinated as part of the catch-up programme against cervical dysplasia.

In our secondary analysis, for routine and younger catch-up study cohorts with outcomes followed from ages 19 to 25 years there were a total of 1,429,486 girls born between 1 September 1992 and 31 August 1997 **(Table 1)**. The proportion of different ethnic groups and proportion belonging to different IMD quintiles was similar between the younger catch-up and the routine vaccinated cohorts. **(Table 2).** At the eligibility point the vaccination rate increased by 8% for two doses between the younger catch-up and routine study cohorts (**Supplementary Table 3**). Overall, all cervical outcomes had lower rates of occurrence over the 6-year follow up per 100,000 persons in girls who were eligible for vaccination as part of the routine programme compared to those eligible for vaccination as part of the catch-up programme (**Table 3**). We did not find a significant discontinuity in any cervical outcomes between the routine and catch-up study cohorts (**Supplementary Figure 3**).

There was no evidence of a change in cancer mortality outcomes in either of our analyses during our follow-up time (**Supplementary Figure 4**).

### Sensitivity analysis

We found no discontinuity in deaths not due to cancer at the eligibility cut-off for the non-vaccinated versus older catch-up or younger catch-up versus routine cohorts (p=0.9) (**Figure 2**, **Table 4**). Pre-determined characteristics (ethnicity and IMD) were continuous across the cut-off between study cohorts indicating that the discontinuity in our cervical outcomes was not influenced by confounding factors (**Supplementary Figure 5**). We found no significant discontinuity in outcomes for cervical cancer or cervical dysplasia using different cut-off values (**Supplementary Table 5**).

**Figure 2:**
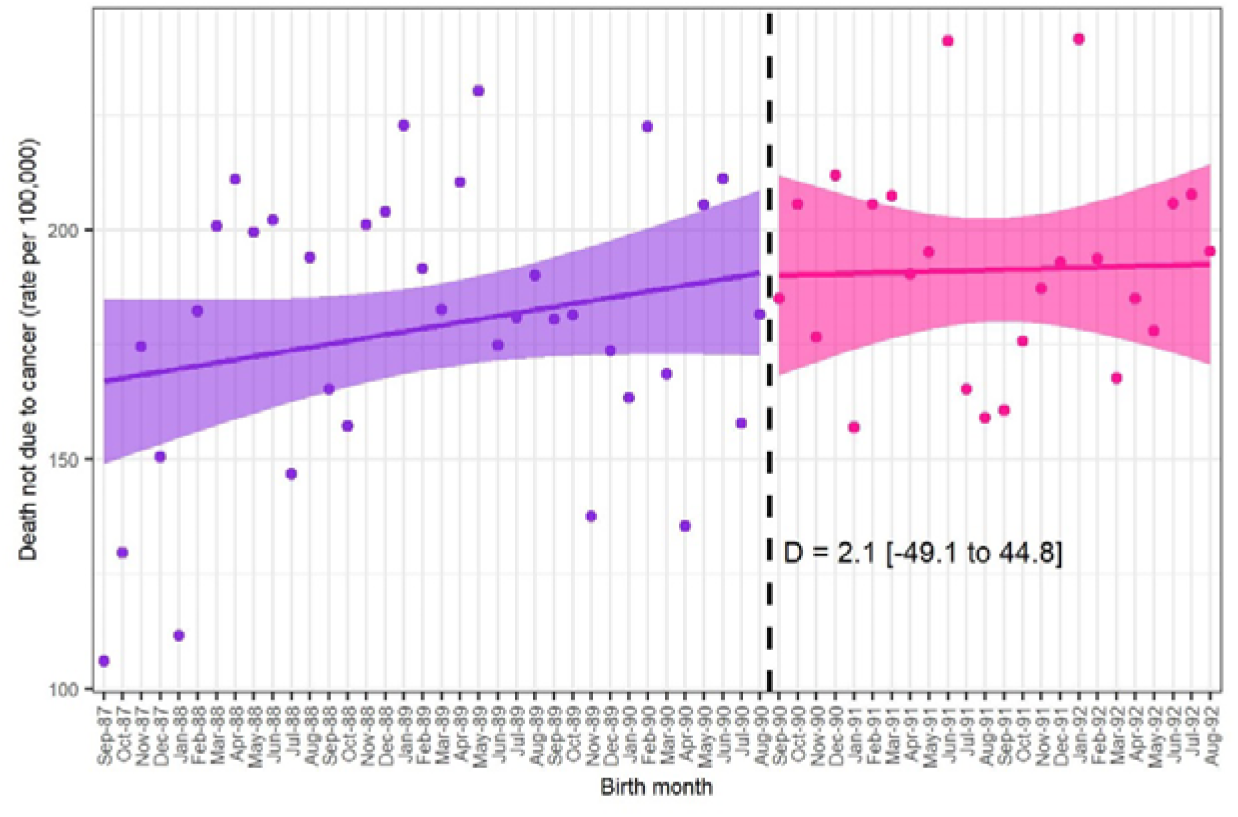
Rate of death not due to cancer across a 7-year follow up period in girls who were eligible and not eligible for HPV vaccination by month of birth. Regression discontinuity design plots for those eligible for HPV vaccination as part of a catch-up programme versus those non-eligible. The rates for those not eligible for the HPV vaccination are shown in purple, and pink for girls eligible for HPV vaccination as part of the catch-up programme. Girls born from 01 September 1990 were eligible for the catch-up vaccination when they were ages 17 to 18 years. The rates are shown for death not due to cancer per 100,000 people. D is the difference in the rate of diagnosis at the discontinuity.

## Discussion

The relationship between HPV vaccination and cervical cancer incidence can be confounded by several factors such as age, lifestyle, and sociodemographic characteristics. Using a quasi-experimental regression discontinuity design approach with population level cohort data we evaluated the vaccine effectiveness between those who were not eligible, received a catch-up vaccination, or were vaccinated as part of the routine programme. We report a clear discontinuity in outcomes in girls who were eligible for vaccination catch up and those who were not. We find that HPV vaccination led to a 31% reduction in the rate of cervical dysplasia and a 75% reduction in the rate of cervical cancer in the oldest women who were vaccinated as part of the catch-up programme compared to the non-vaccinated women over a 7-year follow-up period. We did not find any significant discontinuity in the rates of any outcome for the younger catch-up study cohort compared to the routine study cohort.

Work by Falcaro et al. (2021) in England looked at relative risk of cervical cancer and dysplasia in vaccinated cohorts of young girls compared with non-eligible cohorts [9]. They found an estimated reduction in cervical dysplasia rates ranging from 39% in girls who were offered catch-up vaccination aged 16 to 18 years to 97% in girls who were offered routine vaccination aged 12 to 13 years. For cervical cancer, they found reductions in diagnosis rates ranging from 34% for catch-up at ages 16 to 18, to 87% for routine vaccination at ages 12 to 13 years. Recently published work by Palmer et al. (2024) in Scotland found a reduction in cervical cancer incidence of 74% for women vaccinated with 2 or 3 doses over 14 years of age and no cases of cervical cancer in those vaccinated at ages 12-13 years. They do not report results for cervical dysplasia.

Our results validate the findings of Falcaro et al. (2021) using a different methodology. Our estimate of vaccine effectiveness against cervical dysplasia for the older catch-up study cohort is similar to Falcaro et al. (2021) (31% compared to 39%). There could be slight differences due to the inclusion of 17 to 18-year-olds in our older catch-up study cohort only compared to 16- to 18-year-olds in the oldest cohort in Falcaro et al. (2021), as there is expected to be lower vaccine effectiveness with higher age of administration of the vaccine due to higher background levels of HPV in older girls.

Our estimate of vaccine effectiveness against cervical cancer for the older catch-up study cohort is higher than that of Falcaro et al. (75% compared to 34%) but comparable to that of Palmer et al. (74%). However, our estimate is less precisely with wide confidence intervals. Given incidence of cervical cancer is highest in women aged 30-34 years in the UK, [19] subsequent work using regression discontinuity design should include longer follow-up times when available to assess the real-world effectiveness of the HPV vaccination. Our sensitivity analyses support the validity of using an RDD for the catch-up vs. non vaccinated analysis, with continuity of other covariates across the cut-off and no discontinuity found in a control outcome of death not due to cancer.

We did not find a significant discontinuity at the eligibility cut off for routine vaccination vs. catch-up vaccination for any outcome, however, the difference in vaccination coverage at this point is only 8 percentage points, therefore the power of the RDD to detect a difference in diagnoses or mortality at this cut-off is low. The lack of discontinuity may also be partially a result of herd-protection effects causing a blurring effect across the cut-off time point. We see a decline in the rate of cervical dysplasia as birth month becomes more recent for our main analysis, particularly in the non-vaccine eligible cohort. We also observe this effect in the secondary analysis for cervical dysplasia and cancer diagnoses in the younger catch-up cohort and for cervical dysplasia in the routine vaccinated cohort. This could be related to a decline in screening uptake over time resulting in fewer diagnoses for cervical dysplasia in particular [20]. It could also be related to herd-protection affects for birth cohorts closer to those with higher vaccination rates, increasing vaccination coverage over time due to younger vaccination age for the catch-up birth cohorts in the secondary analysis. This decline results in large differences in crude diagnosis rates between the cohorts, however, crucially, it does not affect the RDD analysis, as decline for these causes would be continuous rather than causing a sudden change at the eligibility cut-offs.

Our analysis did not control for variations in the uptake of cervical screening, as Falcaro et al (2021) did, but nevertheless found similar estimates of vaccination impact on cervical dysplasia. An advantage of the RDD method is that it is not affected by such changes, provided there are no sudden changes that affect women differently either side of the eligibility cut-offs. There were no changes to the policies surrounding cervical screening procedures around the time period when women born close to our eligibility cut-off periods would have been called for their cervical screening around age 25 (∼2015 for the main analysis and ∼2020 for the catch-up vs. routine analysis). The NHS screening programme was launched in 1988 following an increase in mortality rates from cervical cancer in women under the age of 35 years. It initially called women ages 20 to 64 years of age to participate in screening every 3 to 5 years [21]. In 2003 screening age was raised for women to be first called at age 25. Any changes in screening caused by behavioural changes, such as the ‘Jade Goody’ effect (the large increase in cervical screening that followed her death in 2009) will affect all birth cohorts at the cut-off point in the same way and therefore do not need to be accounted for within a RDD.

The detection of cervical disease during 2020 and 2021 was probably reduced by covid disruptions/restrictions interrupting and delaying screening; this may have reduced the rate of screen-detected disease in the oldest birth cohort in our routine study cohort in particular, whose first screen may have been delayed as they reached 25 in 2020 and 2021. The catch-up cohort, and the older unvaccinated, may have also had delayed screening invitations. Importantly, we could not identify any clear reduction in diagnoses, nor changes in average age of diagnoses, associated with the COVID period, so conclude this has not affected our findings.

Our study used a comprehensive, national dataset and a RDD approach to estimate the effectiveness of the HPV vaccine. The RDD reduces the risk of bias due to unobserved factors associated with both the decision to be vaccinated and the risk of adverse outcomes. The fuzzy RDD is comparable to a randomized control trial with imperfect compliance. This approach is not biased by confounding factors in the relationship between the intervention and the disease outcomes as our population will all be affected in the same way, regardless of their relationship to the cut-off. However, the RDD can only be applied in specific circumstances where there is a sudden change in treatment (vaccination) coverage at a change in eligibility for a continuous variable (birth month). Therefore, it cannot for example be used to directly compare outcomes for unvaccinated women with those given routine vaccination and has lower power to detect changes where the change in vaccination coverage is smaller (such as at the switch from catch-up vaccination to routine vaccination).

An important limitation is the relatively short follow-up times for some outcomes. We follow up outcomes for women aged 23 to 30 years, covering two calls for cervical screening. Some outcomes, such as death from cancer, may in future produce detectable differences in incidence between vaccinated and unvaccinated women. Second, our data only include people who responded to the 2011 Census. People who migrated out of the country, chose not to respond, or died before Census are excluded. Whilst the imperfect representativeness may reduce the external validity of this study, it should not affect the internal validity. Survivorship bias is unlikely to be a major issue as the mortality rate in this group is low, and unlikely to differ by eligibility status. Similarly, migration or census non-response is unlikely to be associated with having been eligible the HPV vaccination.

Another limitation of this work is the possibility of ‘spill over’ effects, whereby those who didn’t receive vaccination benefit from the reduction in risk as a consequence of the vaccinated group. In the context of HPV vaccination, HPV is transmitted sexually, and as there is sexual mixing/partnerships between the vaccinated and non-vaccine eligible study cohorts the prevalence of HPV in the non-vaccination eligible study cohort could be reduced.

This would result in our method under-estimating vaccine effectiveness. In addition, the vaccination coverage estimates used are likely to be an underestimate, as shown by seropositivity data [17], and there is no published data on the coverage among girls not eligible for catch-up or routine vaccination. The vaccinated birth cohorts in our study were all offered vaccination with a 3-dose schedule of Cervarix, so our findings are for this vaccine and vaccination schedule. Changes have been made since to the programme’s vaccine and vaccination schedule, most recently to 1-dose of Gardsil [22]. The strong evidence for equivalence of protection from reduced doses suggests schedule changes are unlikely to reduce effectiveness. The use of a 9-valent vaccination should further reduce cervical disease incidence in due course. The RDD approach does not allow direct comparison of routine vaccination and unvaccinated individuals as the birth dates covered are not continuous across these groups.

Whilst we had access to individual-level data for the outcomes, we did not have similar data for vaccination, and had to rely on estimates for each school year and impute vaccination rate by month of birth using these data. This should not cause a large bias to the VE estimate but may have resulted in our confidence intervals being slightly too narrow. The data on vaccination rates may have been underestimates if some vaccinations, such as those given privately, that were not recorded.

Our results have both policy and methodological implications. First, this study demonstrates how successful the HPV vaccination programme has been at reducing cervical disease in young women. Second, it shows that the estimates of the effectiveness of HPV vaccination obtained using a RDD are comparable to those from other studies. This approach can be used in due course to look at the impact of HPV vaccination on other outcomes such as other HPV-related cancers that are rarer and/or are likely to be diagnosed at a later age.

Anal cancer is the second most common HPV related cancer in women, with 80-90% of anal squamous cancers found to be associated with HPV 16/18 infection [23], but is rare in the follow up ages that we study here [24]. This work paves the way for future RDD analysis assessing different cancer outcomes.

## Data Availability

In accordance with NHS Digital's Information Governance requirements, the study data cannot be shared.

## Author contributions

ILW, CRB, and VN conceptualised and designed the study. ILW and CB prepared the study data. ILW and CB performed the statistical analysis, which was quality checked by VN. All authors contributed to interpretation of the findings. ILW and CB wrote the original draft. All authors contributed to review and editing of the manuscript and approved the final manuscript. VN is the guarantor. The corresponding author attests that all listed authors meet authorship criteria and that no others meeting the criteria have been omitted.

