## Supplement for "Evaluating the effectiveness of the HPV vaccination programme in England, using regression discontinuity design"

**Supplementary Material**

### Supplementary Figures

**Supplementary Figure 1: Lexis diagram to show the year of age and vaccination year by birth cohort.**

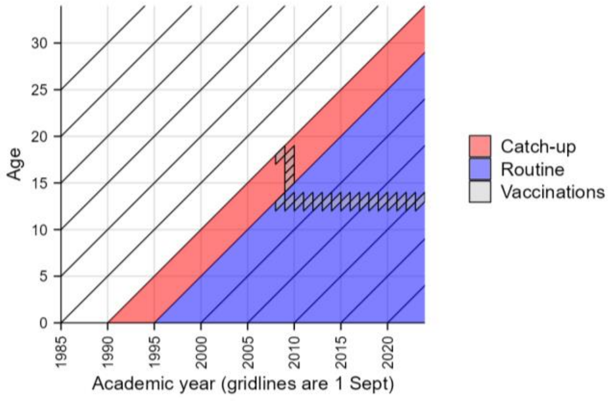


**Supplementary Figure 2: Lexis diagram to show the year of age, vaccination year and age range that outcomes are analysed over for the cohorts used in the analyses and the date ranges for the data available.**

A. Lexis diagram for cohorts used in the main analysis of catch-up vaccination vs. not vaccinated. B. Lexis diagram for cohorts used in the supplementary analysis of routine vaccination vs. catch up vaccination.

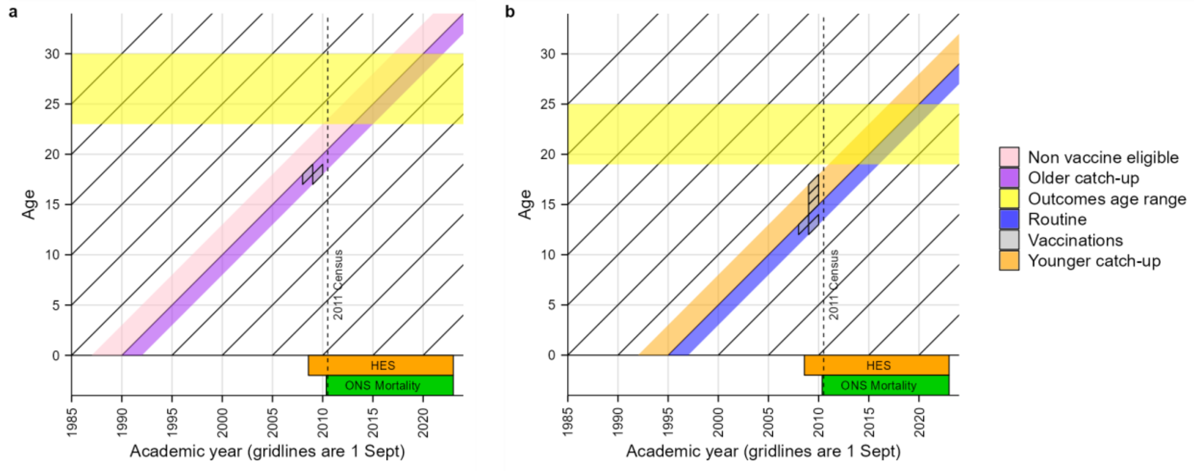

**<u>Supplementary Figure 3:</u> Rate of cervical dysplasia (A) and cervical cancer (B) outcomes across a 6-year follow up period in girls who were eligible for younger catch-up and routine HPV vaccination by month of birth.**

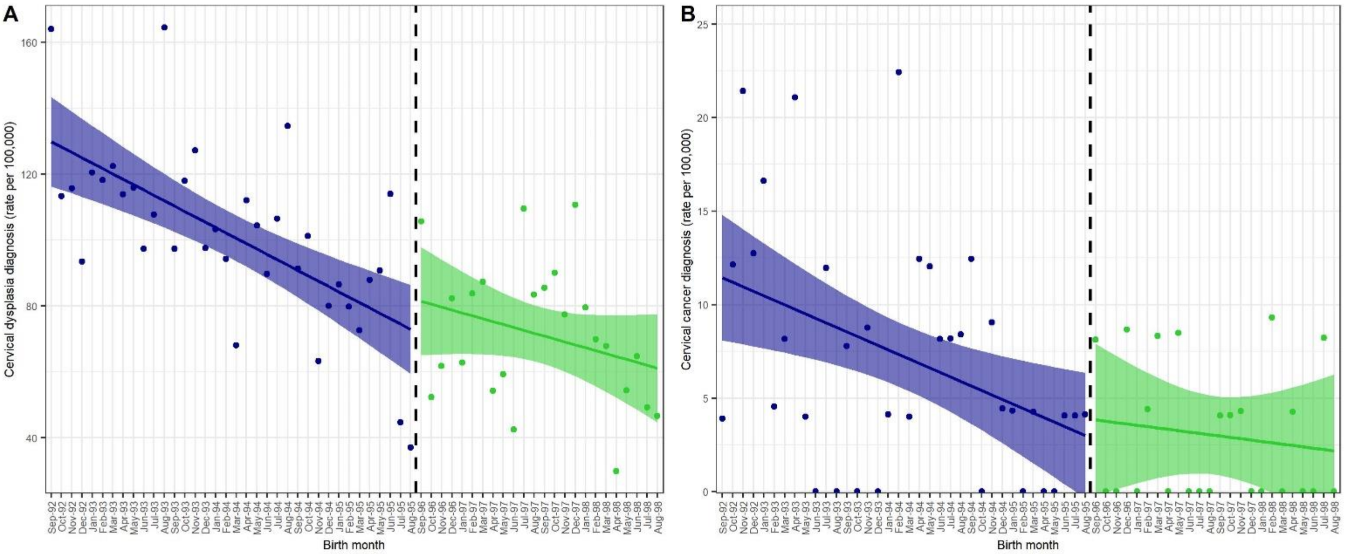


The rates for the youngest 3 birth cohorts eligible for the catch-up HPV vaccination are shown in blue, and green for the first 2 birth cohorts eligible for routine HPV vaccination. Girls born from 1 September 1996 were eligible for the routine vaccination when they were aged 12 to 13 years.

**<u>Supplementary Figure 4:</u> Rate of cervical cancer deaths across a 7-year follow up period by month of birth for women in our main analysis.**

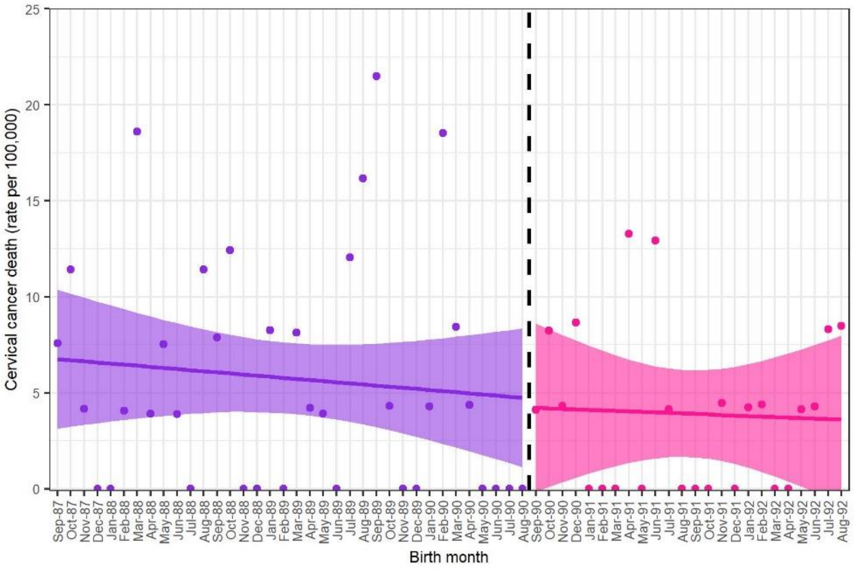


Regression discontinuity design plots for those eligible for HPV vaccination as part of a catch-up programme versus those non-eligible. The rates for those not eligible for the HPV vaccination are shown in purple, and pink for girls eligible for HPV vaccination as part of the catch-up programme. Girls born from 01 September 1990 were eligible for the catch-up vaccination when they were ages 17 to 18 years. The rates are shown for cervical deaths per 100,000 people.

**<u>Supplementary Figure 5: </u>Proportion of girls by ethnic group and IMD group by birth month**

Non-vaccine eligible and older catch-up analysis shown in dark blue (A +B), and routine versus younger catch-up in pink (C + D). Proportion of cohort which belong to the White ethnic group are shown in panels A and C, and the proportion of those belonging to Group 1 of the Index of Multiple Deprivation.

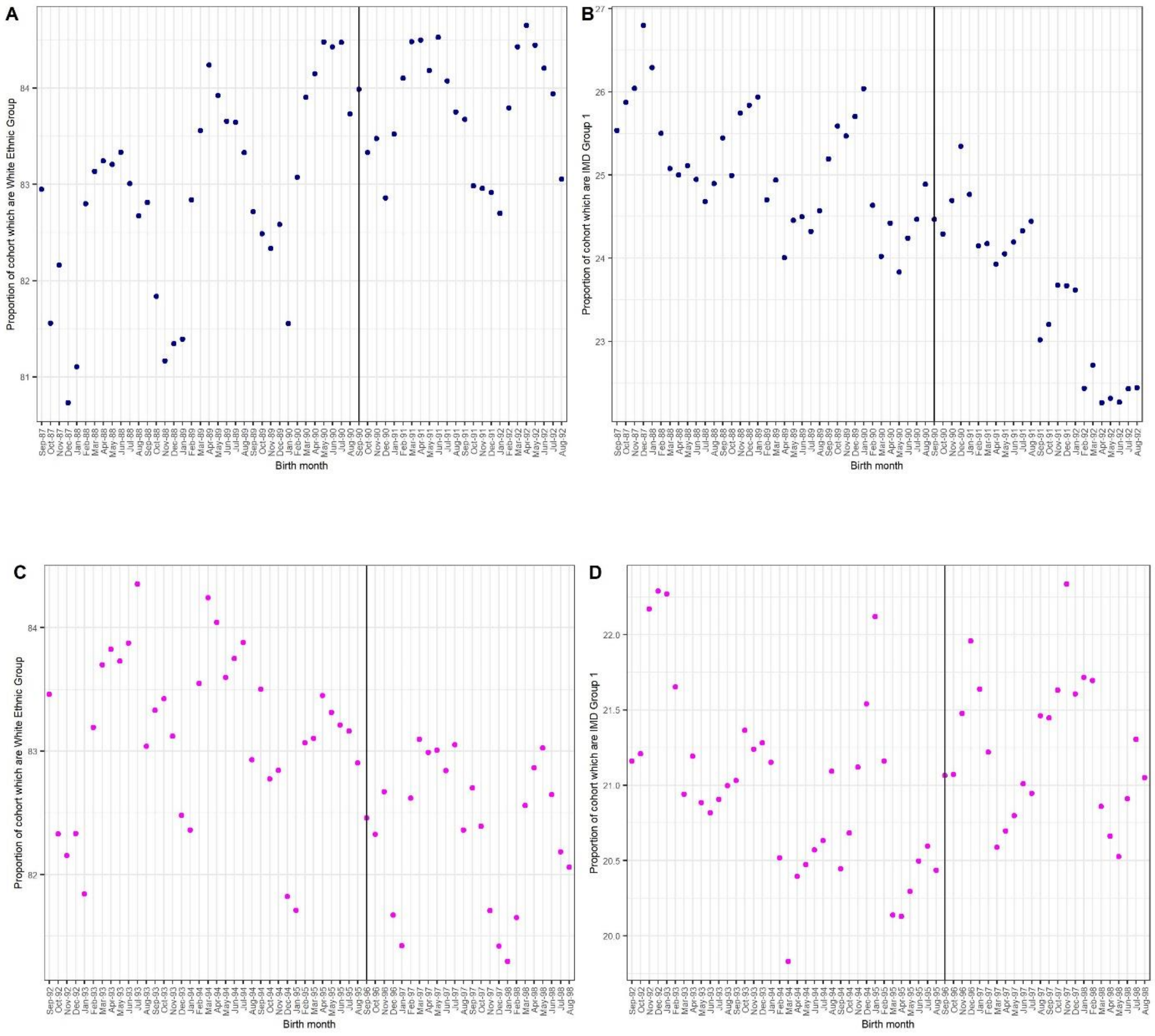

### Supplementary Tables

**Supplementary Table 1: Variables used in the analyses.**

| Variable | Coding | Source |
| --- | --- | --- |
| **Outcomes** | | |
| Death involving cervical cancer | Death registered with cervical cancer (ICD10 codes C530-C539) recorded as contributory or underlying cause of death, where the age at date of death is 19-30. | ONS death registrations |
| Diagnosis of cervical cancer | At least one hospital episode with a primary or secondary diagnosis of cervical cancer (ICD10 codes C530-C539), where the age at the start of the hospital episode is 19-30. | HES |
| Diagnosis of cervical dysplasia | At least one hospital episode with a primary or secondary diagnosis of cervical dysplasia (ICD10 codes N870-N879), where the age at the start of the hospital episode is 19-30. | HES |
| **Exposure** | | |
| Date of birth | Grouped into month of birth. | 2011 Census |

**Supplementary Table 2: Information on cohorts in the main analysis eligible for vaccination and those not based on date of birth. % of vaccinated cohorts was identified from Public Health England (previous publication from Department of Health).**

| Study Cohort | Birth cohort | Vaccinations | Academic year tuned 23 years old | Academic year tuned 30 years old | % vaccinated 1st, 2nd, 3rd doses |
| --- | --- | --- | --- | --- | --- |
| Non vaccine eligible | 1 Sept 1987 - 31 August 1988 | - | 1 September 2010 - 31 August 2011 | 1 September 2017 - 31 August 2018 | 0.01, 0.01. 0,01 |
| Non vaccine eligible | 1 Sept 1988 - 31 August 1989 | - | 1 September 2011 – 31 August 2012 | 1 September 2018 – 31 August 2019 | 0.01, 0.01. 0,01 |
| Non vaccine eligible | 1 Sept 1989 - 31 August 1990 | - | 1 September 2012 – 31 August 2013 | 1 September 2019 – 31 August 2020 | 0.01, 0.01. 0,01 |
| Older catch-up | 1 Sept 1990 - 31 August 1991 | 2008-2009 (year 13/ age 17-18) | 1 September 2013 – 31 August 2014 | 1 September 2020 – 31 August 2021 | 66.1, 59.3, 47.4 |
| Older catch-up | 1 Sept 1991 - 31 August 1992 | 2009-2010 (year 13 / age 17-18) | 1 September 2014 – 31 August 2015 | 1 September 2021 – 31 August 2022 | 55.6, 50.3, 38.9 |

**<u>Supplementary Table 3:</u> Information on cohorts in the secondary analysis eligible for vaccination based on date of birth. % of vaccinated cohorts was identified from Public Health England (previous publication from Department of Health)**

| **Cohort** | **Birth cohort** | **Vaccinations** | **Turned 19** | **Turned 25** | **% vaccinated 1st, 2nd, 3rd** |
|---|---|---|---|---|---|
| Younger catch-up | 1 Sept 1992 - 31 August 1993 | 2009-2010 (Year 12, age 16-17 years) | 1 Sept 2011 - 31 August 2012 | 1 September 2017 – 31 August 2018 | 59.8, **55.9**, 48.1 |
| Younger catch-up | 1 Sept 1993- 31 August 1994 | 2009-2010 (Year 11, age 15-16 years) | 1 Sept 2012 – 31 August 2013 | 1 September 2018–31 August 2019 | 78.4, 75.8, 70.8 |
| Younger catch-up | 1 Sept 1994 - 31 August 1995 | 2009-2010 (Year 10, age 14-15 years) | 1 Sept 2013 – 31 August 2014 | 1 September 2019 – 31 August 2020 | 81.9, 79.6, 75.7 |
| Routine | 1 Sept 1995 - 31 August 1996 | 2008-2009 (Year 8 , age 12-13 years) | 1 Sept 2014 – 31 August 2015 | 1 September 2020 – 31 August 2021 | 89.4, 87.7, 84.4 |
| Routine | 1 Sept 1996 - 31 August 1997 | 2009-2010 (Year 8, age 12-13 years) | 1 Sept 2015 - 31 August 2016 | 1 September 2021 – 31 August 2022 | 85.9, 84.1, 80.9 |

**Supplementary Table 4:** **Total population and rates by birth month for non-vaccinated versus catch-up cohort and catch-up versus routine analysis.**

| Analysis | Birth month | Total (n) | Cervical dysplasia diagnosis (rate per 100,000) | Cervical cancer diagnosis (rate per 100,000) | Cervical cancer mortality (rate per 100,000) | Death not caused by cancer (rate per 100,000) |
|---|---|---|---|---|---|---|
| Main analysis | Sep-87 | 26,411 | 969.3 | 79.5 | 7.6 | 106.0 |
| | Oct-87 | 26,238 | 937.6 | 99.1 | 11.4 | 129.6 |
| | Nov-87 | 24,066 | 993.1 | 87.3 | 4.2 | 174.5 |
| | Dec-87 | 25,246 | 863.5 | 79.2 | 0.0 | 150.5 |
| | Jan-88 | 25,973 | 943.3 | 65.5 | 0.0 | 111.7 |
| | Feb-88 | 24,683 | 879.2 | 89.1 | 4.1 | 182.3 |
| | Mar-88 | 26,883 | 959.7 | 89.3 | 18.6 | 200.9 |
| | Apr-88 | 25,580 | 887.4 | 117.3 | 3.9 | 211.1 |
| | May-88 | 26,562 | 783.1 | 67.8 | 7.5 | 199.5 |
| | Jun-88 | 25,711 | 976.2 | 85.6 | 3.9 | 202.3 |
| | Jul-88 | 25,892 | 876.7 | 73.4 | 0.0 | 146.8 |
| | Aug-88 | 26,291 | 867.2 | 95.1 | 11.4 | 194.0 |
| | Sep-88 | 25,397 | 956.8 | 106.3 | 7.9 | 165.4 |
| | Oct-88 | 24,156 | 898.3 | 103.5 | 12.4 | 157.3 |
| | Nov-88 | 23,356 | 766.4 | 72.8 | 0.0 | 201.2 |
| | Dec-88 | 24,014 | 837.0 | 99.9 | 0.0 | 204.1 |
| | Jan-89 | 24,227 | 862.7 | 107.3 | 8.3 | 222.9 |
| | Feb-89 | 22,434 | 762.2 | 80.2 | 0.0 | 191.7 |
| | Mar-89 | 24,637 | 783.4 | 93.4 | 8.1 | 182.7 |
| | Apr-89 | 23,763 | 816.4 | 54.7 | 4.2 | 210.4 |
| | May-89 | 25,626 | 909.2 | 70.2 | 3.9 | 230.2 |
| | Jun-89 | 24,580 | 817.7 | 93.6 | 0.0 | 174.9 |
| | Jul-89 | 24,878 | 872.3 | 112.6 | 12.1 | 180.9 |
| | Aug-89 | 24,716 | 958.9 | 60.7 | 16.2 | 190.2 |
| | Sep-89 | 23,268 | 795.1 | 68.8 | 21.5 | 180.5 |
| | Oct-89 | 23,148 | 833.8 | 73.4 | 4.3 | 181.4 |
| | Nov-89 | 22,534 | 803.2 | 53.3 | 0.0 | 137.6 |
| | Dec-89 | 23,027 | 881.6 | 65.1 | 0.0 | 173.7 |
| | Jan-90 | 23,251 | 868.8 | 81.7 | 4.3 | 163.4 |
| | Feb-90 | 21,575 | 838.9 | 88.1 | 18.5 | 222.5 |
| | Mar-90 | 23,721 | 843.1 | 71.7 | 8.4 | 168.6 |
| | Apr-90 | 22,870 | 822.0 | 39.4 | 4.4 | 135.6 |
| | May-90 | 24,822 | 809.8 | 24.2 | 0.0 | 205.5 |
| | Jun-90 | 24,158 | 678.9 | 91.1 | 0.0 | 211.1 |
| | Jul-90 | 24,699 | 813.8 | 76.9 | 0.0 | 157.9 |
| | Aug-90 | 24,220 | 858.8 | 90.8 | 0.0 | 181.7 |
| | Sep-90 | 24,319 | 559.2 | 69.9 | 4.1 | 185.0 |
| | Oct-90 | 24,308 | 654.1 | 49.4 | 8.2 | 205.7 |
| | Nov-90 | 23,212 | 611.8 | 47.4 | 4.3 | 176.6 |
| | Dec-90 | 23,123 | 657.4 | 47.6 | 8.7 | 211.9 |

| | | | | | | |
|---|---|---|---|---|---|---|
| | Jan-91 | 23,566 | 577.1 | 34.0 | 0.0 | 157.0 |
| | Feb-91 | 21,399 | 570.1 | 56.1 | 0.0 | 205.6 |
| | Mar-91 | 23,135 | 592.2 | 38.9 | 0.0 | 207.5 |
| | Apr-91 | 22,584 | 611.1 | 79.7 | 13.3 | 190.4 |
| | May-91 | 24,082 | 498.3 | 41.5 | 0.0 | 195.2 |
| | Jun-91 | 23,222 | 611.5 | 51.7 | 12.9 | 241.2 |
| | Jul-91 | 24,185 | 583.0 | 57.9 | 4.1 | 165.4 |
| | Aug-91 | 23,895 | 594.3 | 33.5 | 0.0 | 159.0 |
| | Sep-91 | 24,261 | 618.3 | 24.7 | 0.0 | 160.8 |
| | Oct-91 | 23,890 | 556.7 | 54.4 | 0.0 | 175.8 |
| | Nov-91 | 22,428 | 668.8 | 62.4 | 4.5 | 187.3 |
| | Dec-91 | 22,805 | 605.1 | 26.3 | 0.0 | 192.9 |
| | Jan-92 | 23,586 | 572.4 | 50.9 | 4.2 | 241.7 |
| | Feb-92 | 22,708 | 594.5 | 44.0 | 4.4 | 193.8 |
| | Mar-92 | 23,858 | 591.0 | 29.3 | 0.0 | 167.7 |
| | Apr-92 | 23,230 | 503.7 | 30.1 | 0.0 | 185.1 |
| | May-92 | 24,163 | 554.6 | 74.5 | 4.1 | 178.0 |
| | Jun-92 | 23,323 | 501.7 | 64.3 | 4.3 | 205.8 |
| | Jul-92 | 24,062 | 660.8 | 49.9 | 8.3 | 207.8 |
| | Aug-92 | 23,555 | 522.2 | 80.7 | 8.5 | 195.3 |
| Secondary analysis | Sep-92 | 25,618 | 164.0 | 3.9 | 0.0 | 105.4 |
| | Oct-92 | 24,706 | 113.3 | 12.1 | 0.0 | 117.4 |
| | Nov-92 | 23,342 | 115.7 | 21.4 | 0.0 | 107.1 |
| | Dec-92 | 23,535 | 93.5 | 12.8 | 4.3 | 131.7 |
| | Jan-93 | 24,063 | 120.5 | 16.6 | 0.0 | 87.3 |
| | Feb-93 | 22,006 | 118.2 | 4.5 | 0.0 | 122.7 |
| | Mar-93 | 24,513 | 122.4 | 8.2 | 0.0 | 150.9 |
| | Apr-93 | 23,721 | 113.8 | 21.1 | 0.0 | 109.6 |
| | May-93 | 25,019 | 115.9 | 4.0 | 0.0 | 119.9 |
| | Jun-93 | 24,667 | 97.3 | 0.0 | 0.0 | 145.9 |
| | Jul-93 | 25,064 | 107.7 | 12.0 | 0.0 | 159.6 |
| | Aug-93 | 24,938 | 164.4 | 0.0 | 0.0 | 112.3 |
| | Sep-93 | 25,677 | 97.4 | 7.8 | 0.0 | 148.0 |
| | Oct-93 | 24,583 | 118.0 | 0.0 | 0.0 | 146.4 |
| | Nov-93 | 22,793 | 127.2 | 8.8 | 0.0 | 92.1 |
| | Dec-93 | 23,560 | 97.6 | 0.0 | 0.0 | 123.1 |
| | Jan-94 | 24,207 | 103.3 | 4.1 | 4.1 | 157.0 |
| | Feb-94 | 22,284 | 94.2 | 22.4 | 0.0 | 112.2 |
| | Mar-94 | 24,973 | 68.1 | 4.0 | 0.0 | 92.1 |
| | Apr-94 | 24,100 | 112.0 | 12.5 | 0.0 | 112.0 |
| | May-94 | 24,905 | 104.4 | 12.1 | 4.0 | 112.4 |
| | Jun-94 | 24,510 | 89.8 | 8.2 | 0.0 | 122.4 |
| | Jul-94 | 24,417 | 106.5 | 8.2 | 0.0 | 131.1 |
| | Aug-94 | 23,777 | 134.6 | 8.4 | 0.0 | 117.8 |
| | Sep-94 | 24,098 | 91.3 | 12.5 | 0.0 | 103.7 |
| | Oct-94 | 23,715 | 101.2 | 0.0 | 0.0 | 134.9 |

| | | | | | | |
|---|---|---|---|---|---|---|
| | Nov-94 | 22,122 | 63.3 | 9.0 | 0.0 | 144.7 |
| | Dec-94 | 22,483 | 80.1 | 4.5 | 4.5 | 151.2 |
| | Jan-95 | 23,111 | 86.5 | 4.3 | 0.0 | 99.5 |
| | Feb-95 | 21,294 | 79.8 | 0.0 | 0.0 | 108.0 |
| | Mar-95 | 23,414 | 72.6 | 4.3 | 0.0 | 128.1 |
| | Apr-95 | 22,743 | 87.9 | 0.0 | 0.0 | 145.1 |
| | May-95 | 25,360 | 90.7 | 0.0 | 0.0 | 169.6 |
| | Jun-95 | 24,570 | 114.0 | 4.1 | 0.0 | 122.1 |
| | Jul-95 | 24,637 | 44.7 | 4.1 | 0.0 | 146.1 |
| | Aug-95 | 24,252 | 37.1 | 4.1 | 0.0 | 152.6 |
| | Sep-95 | 24,615 | 105.6 | 8.1 | 0.0 | 117.8 |
| | Oct-95 | 24,821 | 52.4 | 0.0 | 0.0 | 145.0 |
| | Nov-95 | 22,638 | 61.8 | 0.0 | 0.0 | 114.9 |
| | Dec-95 | 23,099 | 82.3 | 8.7 | 0.0 | 112.6 |
| | Jan-96 | 23,902 | 62.8 | 0.0 | 0.0 | 142.3 |
| | Feb-96 | 22,673 | 83.8 | 4.4 | 0.0 | 150.0 |
| | Mar-96 | 24,035 | 87.4 | 8.3 | 0.0 | 112.3 |
| | Apr-96 | 22,096 | 54.3 | 0.0 | 0.0 | 117.7 |
| | May-96 | 23,593 | 59.3 | 8.5 | 0.0 | 114.4 |
| | Jun-96 | 23,528 | 42.5 | 0.0 | 0.0 | 106.3 |
| | Jul-96 | 24,640 | 109.6 | 0.0 | 0.0 | 138.0 |
| | Aug-96 | 23,964 | 83.5 | 0.0 | 0.0 | 104.3 |
| | Sep-96 | 24,558 | 85.5 | 4.1 | 0.0 | 114.0 |
| | Oct-96 | 24,442 | 90.0 | 4.1 | 0.0 | 118.7 |
| | Nov-96 | 23,246 | 77.4 | 4.3 | 0.0 | 103.2 |
| | Dec-96 | 23,494 | 110.7 | 0.0 | 0.0 | 161.7 |
| | Jan-97 | 23,871 | 79.6 | 0.0 | 0.0 | 104.7 |
| | Feb-97 | 21,466 | 69.9 | 9.3 | 0.0 | 97.8 |
| | Mar-97 | 23,573 | 67.9 | 0.0 | 0.0 | 106.1 |
| | Apr-97 | 23,448 | 29.9 | 4.3 | 0.0 | 102.4 |
| | May-97 | 23,907 | 54.4 | 0.0 | 0.0 | 142.2 |
| | Jun-97 | 23,161 | 64.8 | 0.0 | 0.0 | 112.3 |
| | Jul-97 | 24,338 | 49.3 | 8.2 | 0.0 | 131.5 |
| | Aug-97 | 23,601 | 46.6 | 0.0 | 0.0 | 127.1 |

**<u>Supplementary table 5:</u> Sensitivity tests for main analysis changing cut-off month.**

| Outcome | Cut-off birth month | LATE/100,000 (95% CIs) | p-value |
|---|---|---|---|
| Cervical dysplasia | July 1990 | 1886.01 (-17202.77, 20974.79) | 0.85 |
| | Aug 1990 | 5.1 (-980.29, 990.48) | 0.99 |
| | Sept 1990 | -260.21 (-476.92, -43.5) | 0.02 |
| | Oct 1990 | -6756.44 (-7413961.75, 7400448.87) | 1 |
| | Nov 1990 | 17.42 (-1474.88, 1509.72) | 0.98 |
| Cervical cancer | July 1990 | 617.45 (-4911.32, 6146.22) | 0.83 |
| | Aug 1990 | -84.39 (-364.07, 195.29) | 0.55 |
| | Sept 1990 | -87.77 (-170.26, -5.28) | 0.04 |
| | Oct 1990 | 25829.45 (-30061093.46, 30112752.36) | 1 |
| | Nov 1990 | -1.67 (-172.14, 168.8) | 0.98 |
